# Relationship Between Alcohol Consumption and Telecommuting Preference-Practice Mismatch During the COVID-19 Pandemic

**DOI:** 10.1101/2021.12.13.21267711

**Authors:** Chihiro Watanabe, Yusuke Konno, Reiji Yoshimura, Seiichiro Tateishi, Ayako Hino, Mayumi Tsuji, Akira Ogami, Masako Nagata, Keiji Muramatsu, Yoshihisa Fujino, for the CORoNaWork project

**Affiliations:** Department of Psychiatry, University of Occupational and Environmental Health, Japan; Department of Environmental Epidemiology, Institute of Industrial Ecological Sciences, University of Occupational and Environmental Health, Japan; Department of Occupational Medicine, School of Medicine, University of Occupational and Environmental Health, Japan; Department of Mental Health, Institute of Industrial Ecological Sciences, University of Occupational and Environmental Health, Japan; Department of Environmental Health, School of Medicine, University of Occupational and Environmental Health, Japan; Department of Work Systems and Health, Institute of Industrial Ecological Sciences, University of Occupational and Environmental Health, Japan; Department of Occupational Health Practice and Management, Institute of Industrial Ecological Sciences, University of Occupational and Environmental Health, Japan; Department of Public Health, School of Medicine, University of Occupational and Environmental Health, Japan

**Keywords:** Telecommuting, Alcohol Consumption, COVID-19, Japan, Occupational Health

## Abstract

**Introduction:** This study examined the association between increased alcohol consumption and telecommuting, comparing employees who expressed a preference for telecommuting and those who did not.

**Methods:** We conducted an internet monitor survey. Responses from 20,395 of the 33,302 participants were included in the final sample. Participants were asked about their desire for and frequency of telecommuting, and about changes in alcohol consumption under the COVID-19 pandemic. Data were analyzed by logistic regression analysis.

**Results:** Participants who telecommuted despite preferring not to do so reported significantly increased alcohol consumption, as revealed by a multivariate analysis (OR=1.62, 95% CI 1.25-2.12). Participants who expressed a preference for telecommuting showed no such increase.

**Conclusions:** Under the COVID-19 pandemic, telecommuting that involves a mismatch with employee preference for way of working may be a new risk factor for problematic drinking.

## Introduction

With the global outbreak of the new coronavirus, people in many parts of the world are telecommuting. Telecommuting is a way of working that involves working from a location other than one’s regular office or place of business, for example, at home or in a co-working space.^1^ Telecommuting means that one can avoid the flow of people on public transportation and in public spaces during regular commuting, and have fewer contacts with other people at business premises. Therefore, telecommuting is recommend it as a measure to combat the spread of the new coronavirus in many countries, including Japan.^2,3^

Mismatches in telecommuting preferences are more likely to occur in the COVID-19 pandemic than in the past. There are merits and disadvantages to telecommuting. ^4–7^ Merits include increased opportunities to work at one’s own pace, working in one’s own style, lack of stress of commuting, and reduced travel time.

Demerits include difficulties in communication and increased feelings of isolation, lack of space and a suitable desk or chair for work, and difficulty in switching between work and private life. The relative importance of these factors varies among individuals, some of whom will prefer to work from home while others will not. Whereas in the past employees might have had some degree of choice regarding how they worked, today telecommuting may be imposed, regardless of employee preference. This is because telecommuting is recommended as part of the response to the COVID-19 pandemic, for public health reasons. As a result, employees who feel that the disadvantages of telecommuting outweigh the advantages may have to telecommute against their wishes.

People who are required to telecommute despite preferring not to may increase their alcohol intake, as found in a study of Norwegian employees who telecommuted more than 15 hours per week. It was concluded that in response to the instigation of their new style of working, telecommuters drank more to reduce their psychological distress, which included feelings of anxiety and loneliness.^8^ According to a survey by Japan’s Ministry of Finance, spending on alcoholic beverages by working households in December 2020 was higher than in the previous year, when COVID-19 was not prevalent.^9^ By contrast, according to the Japan Foodservice Association (JF) Food Service Industry Market Trend Survey, sales of alcoholic beverages in restaurants in December 2020 was lower than in the year before.^10^

These findings suggest that the consumption of alcohol in Japanese households is on the rise, and this phenomenon is likely to include alcohol consumption by telecommuters. In this internet survey-based study, therefore, we predicted, first, that telecommuting would be associated with increased alcohol consumption in Japan, and second, that this association would be significantly stronger among employees who did not prefer to telecommute.

## Methods

### Study design and participants

This study was done as part of the Collaborative Online Research on the Novel-Coronavirus and Work (CoroNaWork) Project. This project involved an extensive cross-sectional study conducted on the internet in Japan from December 22-26, 2020, investigating the health status of employees during the COVID-19 pandemic. Details of the study protocol are available elsewhere. ^11^ Briefly, data were collected from 33,302 workers who were under contract at the time of survey and who agreed to participate. They were assigned to the study by prefecture, occupation, and gender. Of these, 27,036 were included in this study, after excluding those whose responses were judged to be invalid. Invalid responses included those with extremely short response times, those reporting being less than 140 cm tall or less than 30 kg in weight, and inconsistent responses to multiple identical questions. Furthermore, we excluded workers engaged in jobs where telecommuting is not possible, such as on-site work. Finally, 20,395 workers were included in the analysis. The research was approved by the Ethics Committee of the University of Occupational and Environmental Health, Japan (Reference No. R2-079). Informed consent was obtained via a form on the website.

### Assessment of telecommuting preference and frequency

We used a questionnaire to investigate employees’ attitudes toward and frequency of telecommuting. The following single-item question was used to examine their telecommuting status: “How often do you currently telecommute?” Respondents who answered, “Four or more days a week,” “Two or more days a week,” or “One or more days a week” were categorized as “telecommuting frequency once a week or more;” others were categorized as “telecommuting frequency less than once a week”. We also used the following single-item question to determine participants’ telecommuting preferences: “Would you like to telecommute?” Respondents who answered, “I would rather work at the normal workplace” or “I would prefer to work at the normal workplace as much as possible” were coded as “I do not prefer to telecommute”. Respondents who answered, “I prefer to work from home as much as possible,” “I rather prefer to work from home,” or “I don’t care which,” were classified as “I prefer to telecommute”.

### Assessment of changes in alcohol consumption

We assessed changes in participants’ alcohol consumption during the COVID-19 pandemic. Those who answered “increased” were classified as “with increase” and others as “without increase”.

### Other covariates

The following survey items were considered as potential confounding factors: age, sex, marital status, equivalent income (56-306 JPY/ 318-469 JPY/ 465-1050 JPY), educational background (junior high school/ high school/ university, graduate school, vocational school, junior college), smoking, job type (mainly desk work/ mainly work involving communicating with people), number of employees at the normal workplace (1-29/ 30-99/ 100-999/ ≥1000) and cumulative incidence of COVID-19 in the prefecture. In addition, the cumulative incidence of COVID-19 in the prefecture of residence from the time of the survey to one month before was used as a community-level variable. This latter information was collected from the websites of public institutions.

### Statistical analysis

Age was used as a continuous variable, while other items were presented as categorical variables, using percentages. We conducted a multilevel logistic model nested in the prefectures of residence, with change in alcohol consumption as the dependent variable and preference for telecommuting and frequency of telecommuting as independent variables. To adjust for potential confounders, adjustment for age, sex, marital status, equivalent income, educational level, smoking, alcohol consumption, job type, number of employees at the workplace and cumulative incidence rate of COVID-19 in the prefecture was used as a covariate. We further examined the interaction term between telecommuting frequency and telecommuting preference. All statistical analyses were performed using Stata (Stata Statistical Software: Release 16. College Station, TX: StataCorp LLC.). We considered *p* values less than 0.05 as statistically significant.

## Results

Table 1 presents baseline characteristics of the participants. Of the 20395 participants whose data were used, 5,271 telecommuted at least one day per week. The median age, percentage of married people, and percentage of smokers were similar in the two populations. The proportion of men, the proportion of college graduates, and annual income tended to be higher among those who telecommuted at least one day per week.

**Table 1.**
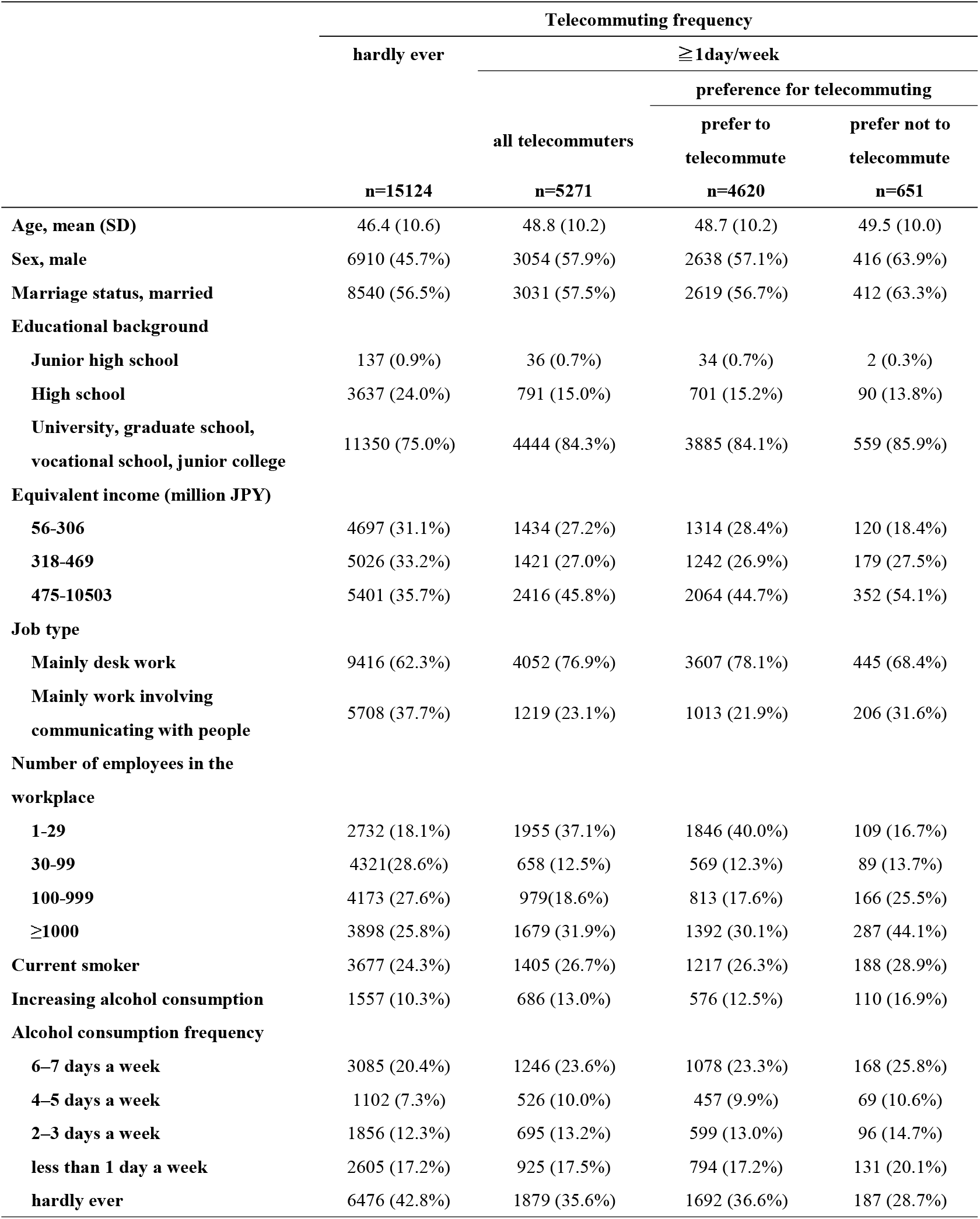
Characteristics of participants by frequency of telecommuting

The odds ratios (ORs) of frequency and preference for telecommuting and alcohol consumption estimated by the logistic model are shown in Table 2. The ratio of increased drinking in those who telecommuted at least once a week was significantly different in univariate analysis compared to those that did little to no telecommuting as a baseline (OR=1.29, 95% CI 1.16-1.43, p<0.001). Multivariate analysis showed similar results (OR=1.21 95%CI1.00-1.29, p=0.001). After adjustment for the interaction term of frequency and preference of telecommuting, there was still a significant difference (OR=1.14, 95% CI 1.00-1.29, p=0.046). The ratio of increased drinking in participants for whom telecommuting was not preferred was not significantly different in univariate analysis using the preferred group as a baseline (OR = 0.97, 95% CI 0.87-1.07, p=0.002). Multivariate analysis showed similar results (OR=0.92 95%CI 0.82-1.03, p=0.001). After adjustment for the interaction term of frequency and preference of telecommuting, there was not a significant difference (OR=0.92, 95% CI 0.82-1.03, p=0.128). The interaction term between frequency of telecommuting and telecommuting preference was significant.

**Table 2.** Relationship between frequency or preference of telecommuting and increasing alcohol consumption

|  | univariate |  |  |  | multivariate* |  |  |  | multivariate** |  |  |  |
| --- | --- | --- | --- | --- | --- | --- | --- | --- | --- | --- | --- | --- |
|  | OR | 95%CI |  | p | OR | 95%CI |  | p | OR | 95%CI |  | p |
| **frequency of telecommuting** |  |  |  |  |  |  |  |  |  |  |  |  |
| telecommuting frequency less than once a week | reference |  |  |  | reference |  |  |  | reference |  |  |  |
| telecommuting frequency once a week or more (19.0%) | 1.29 | 1.16 | 1.43 | <0.001 | 1.21 | 1.08 | 1.36 | 0.001 | 1.14 | 1.00 | 1.29 | 0.046 |
| **preference for telecommuting** |  |  |  |  |  |  |  |  |  |  |  |  |
| prefer telecommuting | reference |  |  |  | reference |  |  |  | reference |  |  |  |
| do not prefer telecommuting (38.9%) | 1.08 | 1.02 | 1.14 | 0.002 | 1.10 | 1.04 | 1.16 | 0.001 | 0.92 | 0.82 | 1.03 | 0.128 |
| **interaction term** |  |  |  |  |  |  |  |  | 1.48 | 1.10 | 1.99 | 0.009 |

\*Multivariate model adjusted for age, sex, marital status, equivalent income, educational level, smoking, alcohol consumption, job type, number of employees at the workplace and cumulative incidence rate of COVID-19 in the prefecture.
\*\* Model further adjusted for interaction term of frequency and preference for telecommuting.

Table 3 shows the odds ratios (ORs) for the frequency of telecommuting and the increase in alcohol consumption for each telecommuting preference category, as estimated by the logistic model. Among participants who did not wish to telecommute, the ratio of increased drinking in the group that telecommuted at least once a week was significantly different in univariate analysis from the group that did little to no telecommuting as a baseline (OR=1.88, 95% CI 1.49-2.38, p<0.001). Multivariate analysis showed similar results (OR=1.62 95%CI 1.25-2.12, p<0.001). There was a significant difference in univariate analysis in the percentage of increased drinking in the group who preferred to telecommute at least once a week with the group who rarely telecommuted as baseline (OR = 1.26, 95% CI 1.13-1.41, p<0.001). However, in multivariate analysis, there was no significant difference (OR=1.10, 95% CI 0.98-1.26, p<0.001).

**Table 3.** Relationship between frequency of telecommuting and increased alcohol consumption for each telecommuting preference

|  | univariate |  |  | multivariate* |  |  |
| --- | --- | --- | --- | --- | --- | --- |
|  | OR | 95%CI | p | OR | 95%CI | p |
| **no preference for telecommuting** |  |  |  |  |  |  |
| telecommuting frequency less than once a week | reference |  |  | reference |  |  |
| telecommuting frequency once a week or more | 1.88 | 1.49 2.38 | <0.001 | 1.62 | 1.25 2.12 | <0.001 |
| **preference for telecommuting** |  |  |  |  |  |  |
| telecommuting frequency less than once a week | reference |  |  | reference |  |  |
| telecommuting frequency once a week or more | 1.26 | 1.13 1.41 | <0.001 | 1.10 | 0.98 1.26 | 0.098 |

\*Multivariate model adjusted for age, sex, marital status, equivalent income, educational level, smoking, alcohol consumption, job type, number of employees at the workplace and cumulative incidence rate of COVID-19 in the prefecture.

## Discussion

Of the employees surveyed, 19% telecommuted at least one day a week. Of these, about 10% did not prefer to telecommute. Our data showed that the frequency of telecommuting more than once a week and the lack of preference for this way of working were associated with increased alcohol consumption. In addition, not preferring to telecommute but telecommuting at least once a week was significantly associated with increased alcohol consumption. By contrast, telecommuting was not associated with an increase in alcohol consumption when telecommuting was preferred. We are aware of no previous study that has examined the association between the desire to telecommute and increased alcohol consumption.

We found that telecommuters had increased alcohol consumption compared to employees who were not telecommuting. Previous studies have also reported that drinking increases among employees who telecommute and have fewer opportunities to go out. In a study of 3,000 telecommuters in the U.S., 32% were more likely to drink during work hours than when they were in their regular workplaces.^12^ Another study found that 18% of Canadians who telecommuted and were less likely to go out drank more frequently.^13^ The causes of increased drinking among telecommuters include social isolation, loneliness, a poor work environment, and disrupted lifestyle.^12,13^ In response to these stressors, employees start to drink more as a coping behavior. In addition, without the inhibitions from monitoring by co-workers and superiors, it is easy to increase alcohol consumption. For this reason, the amount of alcohol consumed by telecommuters is likely to increase. In this study, working from home when this was not the preferred practice was associated with increased alcohol consumption. In contrast, when workers desired to telecommute, telecommuting was not associated with increased alcohol consumption. The results suggest that a mismatch between attitude toward telecommuting and actual working practice, rather than telecommuting itself, is associated with increased alcohol consumption. Whether employees prefer to telecommute depends on which of the merits and demerits of telecommuting has a stronger impact on them. For example, workers who commute long hours may prefer to work from home, while those with no suitable working space at home may prefer not to telecommute. If demerits are perceived more strongly, then telecommuting will not be preferred. Reasons for not wanting to telecommute, such as social isolation, loneliness, poor work environment, and difficulty in establishing a daily rhythm, are consistent with those reported to lead to increased alcohol consumption in previous studies. Drinking alcohol may be a coping behavior against psychological distress caused by social isolation, loneliness, poor work environment, and disordered life rhythm.^14^

The mismatch between preferred and actual telecommuting practices may be a risk factor for inappropriate drinking behavior. It has been pointed out that telecommuting can lead to inadequate on-site supervision,^8^ not only with regard to safety but also the management of employee health. It is difficult for companies to supervise the private life of individual employees, especially their lifestyle. However, inappropriate drinking is associated with health problems including alcohol dependence and liver dysfunction,^15^ along with other problems including presenteeism, decreased productivity, and accident risks.^16–20^ Therefore, deterrence programs and early detection of inappropriate drinking are important issues in occupational health. As telecommuting is expected to become a more widespread and long-term way of working in many companies, placement of employees in ways that are consistent with their preferences for work locations is an important issue.

## Limitations

Several limitations of this study warrant mention. First, because we conducted an internet monitoring survey, the general applicability of the results is uncertain. However, we sought to minimize any bias in the participants by sampling across regions, occupations, and prefectures according to the incidence of infection. Second, we relied on self-reported assessments of alcohol consumption. In general, it has been shown that drinkers tend to underestimate their consumption,^21^ which means that the amount of alcohol consumed might also have been underreported in this study. However, given the anonymous nature of the online survey we believe that underreporting was relatively unlikely. Third, because this is a cross-sectional study, the temporal relationship and direction of causality between telecommuting and increased alcohol consumption is unknown.

## Conclusion

Under the COVID-19 pandemic, telecommuting is recommended as an infection prevention measure. In many cases this has led to the introduction of telecommuting regardless of the preference of the employee, resulting in a mismatch. In this study, 10% of telecommuters did not prefer telecommuting to normal commuting to the workplace. Telecommuting despite no preference for it was significantly associated with increased alcohol consumption. The mismatch between telecommuting-related preferences and imposed practices may be a new risk factor for inappropriate drinking, and therefore an emerging issue in occupational health.

## Data Availability

All data produced in the present study are available upon reasonable request to the authors

## References

1. International Labour Organization. COVID-19: Guidance for labour statistics data collection. Defin Meas Remote Work telework, Work home home-based Work. Published online 2020:14. http://www.ilo.org/global/statistics-and-databases/publications/WCMS_747075/lang--en/index.htm>

2. Advice for the public. Accessed October 25, 2021. http://www.who.int/emergencies/diseases/novel-coronavirus-2019/advice-for-public>

3. “New lifestyle” assuming a new coronavirus | Ministry of Health, Labor and Welfare. Accessed October 25, 2021. https://www.mhlw.go.jp/stf/seisakunitsuite/bunya/0000121431_newlifestyle.html>

4. Montreuil S, Lippel K. Telework and occupational health: a Quebec empirical study and regulatory implications. Saf Sci. 2003;41(4):339-358. doi:10.1016/S0925-7535(02)00042-5

5. Mann S, Holdsworth L. The psychological impact of teleworking: Stress, emotions and health. New Technol Work Employ. 2003;18(3):196-211. doi:10.1111/1468-005X.00121

6. Xiao Y, Becerik-Gerber B, Lucas G, Roll SC. Impacts of working from home during COVID-19 pandemic on physical and mental well-being of office workstation users. J Occup Environ Med. 2021;63(3):181-190. doi:10.1097/JOM.0000000000002097

7. Nagata T, Ito D, Nagata M, et al. Anticipated health effects and proposed countermeasures following the immediate introduction of telework in response to the spread of COVID-19: The findings of a rapid health impact assessment in Japan. J Occup Health. 2021;63(1):e12198. doi:10.1002/1348-9585.12198

8. Nielsen MB, Christensen JO, Knardahl S. Working at home and alcohol use. Published online 2021. doi:10.1016/j.abrep.2021.100377

9. Statistics Bureau household survey results. Accessed October 25, 2021. <https://www.stat.go.jp/data/kakei/2.html#new>

10. Japan Food Service Association. Accessed October 25, 2021. http://www.jfnet.or.jp/data/data_c.html

11. Fujino Y, Ishimaru T, Eguchi H, et al. Protocol for a nationwide internet-based health survey of workers during the COVID-19 pandemic in 2020. J UOEH. 2021;43(2):217-225. doi:10.7888/JUOEH.43.217

12. American Addiction Centers Resourse. Drinking alcohol when working from home. Https://WwwAlcoholOrg/Guides/Work-From-Home-Drinking/. Published online 2020. <https://www.alcohol.org/guides/work-from-home-drinking/>

13. Canadian Centre on Substance Use and Addiction. COVID-19 and increased alcohol consumption. 2020;(April). <https://www.ccsa.ca/covid-19-and-increased-alcohol-consumption-nanos-poll-su> mmary-report

14. Khantzian EJ. The self-medication hypothesis of addictive disorders: focus on heroin and cocaine dependence. https://doi.org/101176/ajp142111259. 2006;142(11):p1259-1264. doi:10.1176/AJP.142.11.1259

15. EJ E, DA F. In the Clinic. Alcohol use. Ann Intern Med. 2016;164(1):ITC1-ITC16. doi:10.7326/AITC201601050

16. Jørgensen MB, Pedersen J, Thygesen LC, et al. Alcohol consumption and labour market participation: a prospective cohort study of transitions between work, unemployment, sickness absence, and social benefits. Eur J Epidemiol. 2019;34(4):397-407. doi:10.1007/s10654-018-0476-7

17. Böckerman P, Hyytinen A, Maczulskij T. Alcohol consumption and long-term labor market outcomes. Published online 2015. doi:10.1002/hec.3290

18. Macdonald Z, Shields MA. The Impact of Alcohol Consumption on Occupational Attainment in England. Published online 1998.

19. MacDonald Z, Shields MA. Does problem drinking affect employment? Evidence from England. Health Econ. 2004;13(2):139-155. doi:10.1002/HEC.816

20. Dooley D, Catalano R, Hough R. Unemployment and alcohol disorder in 1910 and 1990: Drift versus social causation. J Occup Organ Psychol. 1992;65(4):277-290. doi:10.1111/J.2044-8325.1992.TB00505.X

21. Knibbe RA, Bloomfield K. Alcohol consumption estimates in surveys in Europe: Comparability and sensitivity for gender differences. Subst Abus 2001 221. 2001;22(1):23-38. doi:10.1023/A:1026419825284

